# Predictive value of *SLCO1B1* c.521T>C polymorphism on observed changes in the treatment of 1136 statin-users

**DOI:** 10.1101/2022.08.29.22279331

**Authors:** ME Jansen, T Rigter, TMC Fleur, PC Souverein, WMM Verschuren, SJ Vijverberg, JJ Swen, W Rodenburg, MC Cornel

**Affiliations:** Section Community Genetics, Department of Clinical Genetics and Amsterdam Public Health research institute, Amsterdam UMC, Vrije Universiteit Amsterdam, Amsterdam, The Netherlands; Centre for Health Protection, National Institute for Public Health and the Environment, Bilthoven, The Netherlands; Division of Pharmacoepidemiology & Clinical Pharmacology, Utrecht Institute for Pharmaceutical Sciences, Utrecht University, Utrecht, The Netherlands; Centre for Nutrition, Prevention and Health Services, National Institute for Public Health and the Environment, Bilthoven, The Netherlands; Department of Respiratory Medicine, Amsterdam Public Health Institute, Amsterdam UMC, University of Amsterdam, Amsterdam, The Netherlands; Department of Clinical Pharmacy & Toxicology, Leiden University Medical Centre, Leiden, The Netherlands; Julius Center for Health Sciences and Primary Care, University Medical Center Utrecht, Utrecht, The Netherlands

**Keywords:** Statins, pharmacogenomics, primary care, screening, adverse drug reactions

## Abstract

**Purpose:** Pharmacogenomic testing is a method to prevent adverse drug reactions. Pharmacogenomics could be relevant to optimize statin treatment, by identifying patients at high risk for adverse drug reactions. We aim to investigate the clinical validity and utility of pre-emptive pharmacogenomics screening in primary care, with *SLCO1B1* c.521T>C as a risk factor for statin induced adverse drug reactions.

**Methods:** The focus was on changes in therapy as a proxy for adverse drug reactions observed in statin-users in a population-based Dutch cohort. In total 1136 statin users were retrospectively genotyped for the *SLCO1B1* c.521T>C polymorphism (rs4149056) and information on their statin dispensing was evaluated as a cross-sectional research.

**Results:** Approximately half of the included participants discontinued or switched their statin treatment within three years. In our analyses we could not confirm an association between the *SLCO1B1* c.521T>C genotype and any change in statin therapy or arriving at a stable dose sooner in primary care.

**Conclusion:** To be able to evaluate the predictive values of *SLCO1B1* c.521T>C genotype on adverse drug reactions from statins, prospective data collection of actual adverse drug reactions and reasons to change statin treatment should be facilitated.

## Introduction

Pharmacogenomic testing (PGx) is a method to prevent adverse drug reactions (ADRs). PGx could be relevant to optimize statin treatment [1, 2]. Statins reduce low-density-lipoprotein cholesterol (LDL-c) levels and lower the risk for cardiovascular events [3]. Statins are prescribed to millions of patients and are usually well-tolerated [4], however statin related myopathy (SRM) is estimated to manifest in 10-25% of patients in clinical practice [5-9]. Approximately 30% of patients discontinue treatment without consulting their physician within their first year, most likely due to ADRs such as mild SRM [5, 6, 9]. Patients who do not consult their physician, might be at increased risk for cardiovascular events since they do not get alternative treatment, with inherent medical and economic consequences [10, 11]. By preemptive screening PGx related to statins, SRM and subsequent non-compliance can be reduced and patients can receive a more effective treatment, resulting in better health outcomes. Therefore we studied the influence of one PGx-variant on SRM.

SRM ranges from mild myalgia (skeletal muscle pain without evidence of muscle degradation) to rhabdomyolysis (severe skeletal muscle damage with acute kidney injury) [12]. The risk of developing SRM depends on several factors. The risk is higher for more lipophilic statins, such as simvastatin, especially at a high dose [5, 13]. Secondly, demographics and life style factors contribute to this risk, such as higher age, lower body mass index (BMI), and being female [5, 6, 9]. Furthermore, the solute carrier organic anion transporter family member 1B1 (*SLCO1B1*) c.521T>C polymorphism (rs4149056) is associated with SRM [5, 9, 14-17]. To prevent SRM, the Clinical Pharmacogenetics Implementation Consortium (CPIC) and the Dutch Pharmacogenetics Working Group (DPWG) provide PGx-guidelines for dosing simvastatin and atorvastatin based on the *SLCO1B1* c.521T>C SNP [12, 18-20]. In the CPIC guideline on *SLCO1B1* and SRM it is stated theoretically 30 patients need to be genotyped to prevent one ADR.

In a meta-analysis, Xiang et al. (2018) concluded that the association between SLCO1B1 and SRM is not consistent [21]. While the overall odds ratios seems to indicate there is an association with SRM, especially for simvastatin, significant results were not confirmed in the included fourteen studies. The genome-wide study of the SEARCH Collaborative Group in 2008 was the first to report a strong association between the single nucleotide polymorphism (SNP) c.521T>C (rs4149056) in *SLCO1B1* and SRM in patients treated with 80 mg simvastatin daily.[22] The risk for myopathy in CC homozygotes was higher than in TT homozygotes (odds ratio (OR) 16.9; 95% CI 4.7 – 61.1; 85 cases and 90 controls). Link et al. (2008) replicated this result in participants of the Heart Protection Study taking 40 mg simvastatin (per C allele RR 2.6; 95% CI 1.3 – 5.0; 21 cases and 16,643 controls) [22]. Four other studies have confirmed this association between *SLCO1B1* and SRM with simvastatin [23-26].

The association of SRM with the *SLCO1B1* c.521T>C genotype has not been established for atorvastatin [23, 24, 26-28]. Only Puccetti *et al*. (2010) have reported a statistically significantly increased risk in 46 patients with familial hypercholesterolemia taking 20-40 mg atorvastatin per day [29]. When multiple statins were combined a statistically significant increased risk was found in five studies [23-26, 30]. However, Linde *et al*. (2010) and Brunham *et al*. (2012) did not report a statistically significant increased risk in a case-control study of respectively 27 and 25 cases and 19 and 83 controls [27, 31]. These two studies are in line with the results of Hubáček *et al*. (2015) in a Czech population of 3294 patients receiving 10-20 mg simvastatin or atorvastatin per day, who also did not find a statistical significant association [7].

The clinical relevance of PGx for statins remains debated because of the small effect sizes of other studies than the SEARCH Collaborative Group and number of patients that are expected to benefit, while the prevalence of the heterozygous *SLCO1B1* c.521T>C genotype in the general population is estimated to between 14 – 22% [15, 22, 32]. The prevalence of the relevant PGx variant, together with the large group of patients receiving statin prescriptions, might make even slight risk reductions relevant for practice. To be able to evaluate the contribution PGx delivers to effective and safe drug therapy, information on clinical validity and utility is needed. Clinical validity and utility of genetic tests should be assessed before implementation [33]. Clinical validity focuses on the discriminative ability of a test [33]. Clinical utility implies that a genetic test should impact on health outcomes in a relevant way in clinical practice [34-36]. Therefore, the net benefit of health outcomes also depends on the clinical context, such as when and where is PGx applied (figure 1) [35, 37]. PGx has already proven clinically useful in secondary health care [38, 39]. However, as the CPIC and DPWG guidelines illustrate, PGx could also contribute to treatments in primary health care, such as statins [12, 40].

**Fig. 1.**
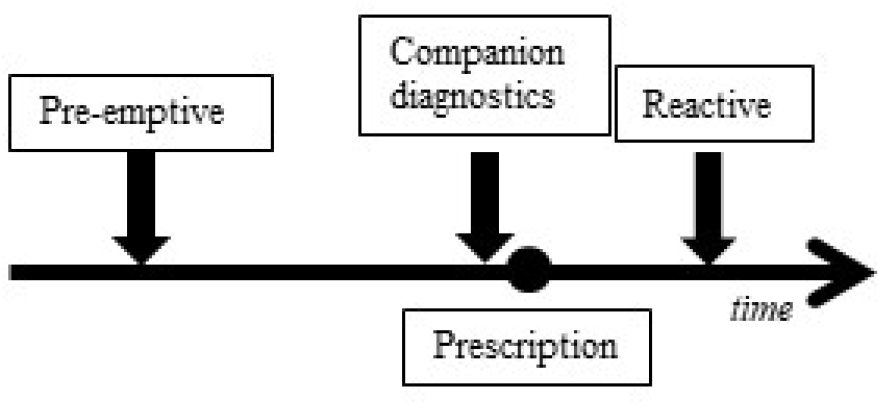
Pre-emptive testing is unrelated to a specific treatment and performed independent of a medical indication. Companion diagnostic testing is performed at the time of prescribing to choose the right drug or dosage. Reactive testing occurs after a patient has started a drug treatment. The aim of reactive testing is to find an explanation for side effects and improve an existing drug therapy.

## Aim

By identifying the *SLCO1B1* c.521T>C SNP pre-emptively or as companion diagnostics (CDx) (figure 1), statin therapy can become more efficacious while SRM is prevented. However, the clinical validity and utility have not been established. The starting point of clinical validity is whether the *SLCO1B1* risk genotype is associated with any ADR. Therefore we aimed to analyze if the *SLCO1B1* c.521T>C is associated with any change in statin therapy in participants in a population based Dutch cohort study.

## Methods

### Study population

The Doetinchem Cohort Study (DCS) is an ongoing longitudinal population-based cohort of randomly selected Dutch inhabitants of Doetinchem, aged 20-59 years at baseline (1987-1991) [41, 42]. Participants have been re-examined once every five years since the baseline measurements. Data from the Doetinchem cohort was linked to the Out-patient Pharmacy Database of the PHARMO Database Network. This database comprises GP or specialist prescribed healthcare products dispensed by the out-patient pharmacy. The dispensing records include information on type of product, date, strength, dosage regimen, quantity, route of administration, prescriber specialty and costs. Informed consent for linkage between data of the Doetinchem cohort and the PHARMO Database Network was acquired before linkage was conducted [42].

Participants were included in this study when a blood sample was available and when they had at least one dispensing of simvastatin or atorvastatin between 01-07-1998 and 31-08-2015. While data was available from 1998 onwards, the starting date for our study (01-07-1998) was chosen to increase reliability on first start of statin therapy, because it was unknown whether patients were using statins before 01-01-1998. Patients with a dispensing before 01-07-1998 were excluded from the study population.

### Follow-up

After inclusion, participants were followed from the date of the first statin dispensing until 1) a follow-up period of three years was achieved, 2) the final date for the current analyses was reached (31-08-2015) within three years after follow-up, 3) the patient died or 4) loss to follow-up. A period of three years of follow-up was chosen because the risk for SRM is the highest during the first years of statin therapy [24].

### Genotyping

Blood samples were taken during every (re-)examination and cryopreserved. DNA was extracted from buffy coats by a salting out method [43]. Using the QuantStudio™ 12 K Flex Real-time PCR system (Life Technologies, Bleiswijk, the Netherlands) with the PGx Express Panel array (ThermoFischer Scientific), DNA was analyzed for the SNP c.521T>C in *SLCO1B1* according to the manufacturer’s protocol between September – November 2017.

### Outcomes

We used proxies to assess the dispensing policy and the number of SRM. The first outcome was *the difference in dose change*. This was measured by comparing the dose category of the first dispensing to dose category of the last dispensing. The dose categories of simvastatin (ATC code C10AA01) were 10, 20 and 40 mg and atorvastatin (ATC code C10AA05) were 5, 10, 20, 40, and 80 mg based on the defined daily dose equivalent (DDD-E).

The second outcome was *any change in drug use*. This was categorized by discontinuation and switching. Discontinuation was defined as no new statins dispensed within 90 days after the theoretical end date of the last dispensing and no other cholesterol-lowering drug within 90 days after the end of the last dispensing or no further dispensing issued for any cholesterol-lowering drug and more than 90 days available to the right censoring date. Switching was defined as no new statins dispensed within 90 days after the theoretical end date of the last dispensing and another cholesterol-lowering drug within 90 days after the theoretical end date of the last dispensing [44, 45]. Switching was specified by changing from simvastatin or atorvastatin to another statin (simvastatin, atorvastatin, rosuvastatin, pravastatin or fluvastatin) or cholesterol-lowering drug (acipimox, ezetimibe, bezafibrate, ciprofibrate, fenofibrate, gemfibrozil, colesevelam, cholestyramine, nicotinic acid, docosapentaenoic acid or eicosapentaenoic acid) [46].

The third outcome was *the difference in time to establish stable dosing*, defined as the time until three times successively the same dose was dispensed based on DDD-E. After this moment, the dosing regimen was assumed to be stable. The time to establish stable dosing was used as a measure for health impact, i.e. when a patient has a stable dosing regimen sooner they are assumed to have less SRM and need less health care visits.

### Covariables

#### Potential confounders

Potential confounders were identified before analysis and grouped into three categories: lifestyle factors (cigarette smoking, physical activity, BMI), biological factors (age, sex, diabetes mellitus, systolic blood pressure, HDL level) and drug-related factors (starting dose, concomitant drug use) [9, 12, 47, 48]. Specific concomitant drugs assessed were: amiodarone, diltiazem, verapamil, fluconazole, ketoconazole, clarithromycin, erythromycin, cyclosporine, gemfibrozil and HIV protease inhibitors, as all inhibit statin metabolism through CYP3A4 or OATP1B1 proteins.

#### Effect modifier

The type of initial statin was assumed an effect modifier, based on the different pathways in which simvastatin and atorvastatin are metabolized in the body. Therefore groups were stratified according to the statin at first dispensing.

### Statistical analysis

The Hardy-Weinberg equilibrium was calculated to confirm that the allele frequency in the study population was constant and in balance by using a Chi-squared test. The genetic model was co-dominant, where each mutant allele contributes to the amount that a patient is affected, in line with the OR per C-allele reported by Link *et al*. (2008) [22]. Sample size for a power of 80% was calculated (Supplementary Material 1). Based on stratification for one variable and a dropout rate of 10%, sample size has to be at least 176 patients, approximately 44 per compared group. The outcome of the study used for the power calculation was myopathy and the daily dose was 80 mg of simvastatin. We expect that our sample needs to be bigger to correct for the regular lower dose and the less directly measured outcomes.

Baseline characteristics of the different genotype groups were compared by linear regression for continuous variables and Chi-squared tests for categorical variables to notice significant deviations. *The difference in time to establish the stable dosing* was analyzed with Cox proportional hazard analysis. *The difference in dose change* and *any change* ***in*** *drug use* (discontinuation and switch) were analyzed with logistic regression. Stratification was limited to initial statin, because stratification into smaller subgroups could result in not having enough power. All analyses were conducted with SPSS software (version 24.0). A p-value ≤ 0.05 was considered to be statistically significant. ORs, HRs, and test characteristics were calculated separately for simvastatin and atorvastatin.

## Results

### Population sample

After applying the inclusion criteria, 2,226 statin users could be linked between the PHARMO Database Network and 5,683 DCS-participants (39.2%). For the current study, measurements for DCS-participants were considered eligible, if a participant has participated in at least three rounds of measurements. Therefore, another 1,067 participants were excluded, resulting in 1,159 included statin users (52.1%). Of the included statin users, 1,136 (98.0%) were successfully genotyped for *SLCO1B1*. In table 1 the baseline characteristics of our study population are shown. At baseline, 928 (81.7%) participants were using simvastatin and 208 (18.3%) atorvastatin. Start dose of simvastatin was lower in women compared to men (DDD eq. at baseline median 0.7 versus 1.3; p=0.054). The genotypes were in Hardy-Weinberg equilibrium (χ^2^ = 0.009; p = 0.924) (Supplementary Material 2).

**Table 1.** Baseline characteristics of 1136 statin users from the Doetinchem cohort, categorized by *SLCO1B1* c.521T>C genotype.

| *SLCO1B1* genotype | TT (n=789) | TC (n=316) | CC (n=31) | Total |
| --- | --- | --- | --- | --- |
| Sex |  |  |  |  |
| Male, % | 51.1 | 51.3 | 38.7 | 50.8 |
| Female, % | 48.9 | 48.7 | 61.3 | 49.2 |
| Age (years), mean (±SD) | 62.5 (9.1) | 62.8 (9.1) | 61.8 (9.4) | 62.6 (9.1) |
| BMI (kg/m<sup>2</sup>), mean (±SD) | 27.8 (4.5) | 27.4 (4.5) | 28.3 (4.4) | 27.7 (4.6) |
| Diastolic blood pressure (mmHg), mean (±SD) | 83.5 (10.9) | 83.6 (10.4) | 82.5 (11.0) | 83.5 (10.7) |
| Systolic blood pressure (mmHg), mean (±SD) | 136.9 (18.4) | 136.0 (18.0) | 136.5 (21.0) | 136.5 (18.4) |
| Total cholesterol (mmol/L), mean (±SD) | 5.9 (1.2) | 5.9 (1.2) | 6.2 (1.3) | 5.9 (1.2) |
| Smoking |  |  |  |  |
| Current, % | 27.9 | 25.2 | 32.3 | 27.3 |
| Past, % | 45.9 | 45.5 | 48.4 | 45.8 |
| Never, % | 26.2 | 29.3 | 19.4 | 26.9 |
| Statin dispensed |  |  |  |  |
| Simvastatin, % | 83.8 | 76.6 | 80.6 | 81.7 |
| Atorvastatin, % | 16.2 | 23.4 | 19.4 | 18.3 |
| Start dose simvastatin (mg) |  |  |  |  |
| 10 mg, % | 11.0 | 8.8 | 12.0 | 10.4 |
| 20 mg, % | 37.7 | 45.8 | 52.0 | 40.2 |
| 40 mg, % | 51.3 | 45.4 | 36.0 | 49.3 |
| Start dose atorvastatin (mg) |  |  |  |  |
| 5 mg, % | 0.3 | 0.0 | 0.0 | 0.2 |
| 10 mg, % | 57.0 | 54.1 | 66.7 | 56.3 |
| 20 mg, % | 33.6 | 36.5 | 33.3 | 34.6 |
| 40 mg, % | 6.3 | 8.1 | 0.0 | 6.7 |
| 80 mg, % | 1.6 | 1.4 | 0.0 | 1.4 |

### Dose change

Among the simvastatin users, 31 (3.3%) participants changed to a lower dose at the end of follow-up compared to their start dose, and 8 (3.8%) patients in the atorvastatin group. There were no statistically significant differences between the TT and TC/CC genotype groups (Table 2).

**Table 2.** The association between *SLCO1B1* c.521T>C genotype and discontinuation, switch, time to establish a stable dose, by statin type.

|  |  | Simvastatin |  | Atorvastatin |  |
| --- | --- | --- | --- | --- | --- |
| Dose change | Event (%) | Crude OR (95% CI) | Adjusted OR<sup>a</sup> (95% CI) | Crude OR (95% CI) | Adjusted OR<sup>b</sup> (95% CI) |
| TT | 30 (4.1) |  |  |  |  |
| TC/CC | 9 (3.0) | 0.50 (0.19-1.31) | 0.42 (0.15-1.16) | 1.72 (0.42-7.11) | 1.97 (0.43-9.04) |
| Total | 39 (3.4) |  |  |  |  |
| Change (discontinuation or switch) | Event (%) | Crude OR (95% CI) | Adjusted OR<sup>a</sup> (95% CI) | Crude OR (95% CI) | Adjusted OR<sup>a</sup> (95% CI) |
| TT | 529 (67.1) |  |  |  |  |
| TC/CC | 223 (64.3) | 0.93 (0.69-1.25) | 0.80 (0.58-1.11) | 0.58 (0.31-1.09) | 0.57 (0.27-1.18) |
| Total | 752 (66.2) |  |  |  |  |
| Time to establish stable dosing regimen | Median (IQR), days | Crude OR (95% CI) | Adjusted OR<sup>a</sup> (95% CI) | Crude OR (95% CI) | Adjusted OR<sup>a</sup> (95% CI) |
| TT | 90 (97) |  |  |  |  |
| TC/CC | 89 (108) | 1.02 (0.87-1.21) | 1.06 (0.89-1.26) | 0.84 (0.61-1.14) | 0.82 (0.57-1.16) |
| Total | 90 (97) |  |  |  |  |

OR = Odds Ratio; 95% CI = 95% confidence interval Adjusted for: a. Age, Sex, Diabetes Mellitus, Systolic blood pressure, HDL level, BMI, cigarette smoking, physical activity, starting dose, concomitant drug use; b. because of small sample size, a limited number of confounders could be added in the model: Age, Sex, Diabetes Mellitus, Systolic blood pressure, HDL level.

### Any change in drug use: discontinuation and switching

Within the 3 years follow-up 535 (47.1%) of the 1136 statin users, discontinued their treatment and 217 (19.1%) switched treatment. The discontinuation proportion between simvastatin and atorvastatin users was comparable (47.2% vs. 46.6%, p=0.812), while more atorvastatin users switched their treatment compared to simvastatin users (27.4% vs 17.2%, p=0.001). Among simvastatin users, a TC/CC genotype was not significantly associated with any change in statin treatment (OR = 0.93, 95% CI 0.69 – 1.25) compared to a TT genotype. Users of atorvastatin with a TC/CC genotype did not change their treatment more often compared to patient with a TT genotype either (OR = 0.58, 95% CI 0.31 – 1.09) (table 2).

### Test characteristics for SRM

There was no association between *SLCO1B1* 521T>C and switch or discontinuation, so the test characteristics of genotyping for *SLCO1B1* 521T>C were not calculated: sensitivity, specificity, positive predictive value, negative predictive value.

### Time to establish stable dosing

The median time to establish stable dosing was 90 days (IQR 97 days) days. Among *SLCO1B1* TT genotype the median time was 90 days (IQR 97 days), and among *SLCO1B1* TC/CC genotypes 89 days (IQR 108). For simvastatin and atorvastatin users it took respectively a median number of days of 89 (IQR 96 days) and 92 (IQR 108 days) until successively three times the same dosing was dispensed. The time to establishing a stable dose was not significantly different between TT genotype and TC/CC genotypes, both for simvastatin users and for atorvastatin users (table 2).

## Discussion

In this study we aimed to investigate the clinical validity and clinical utility of pre-emptive pharmacogenomics *SLCO1B1* c.521T>C screening in primary care. The starting point of clinical validity is whether the *SLCO1B1* risk genotype is associated with any change in statin therapy. In our cohort study 30.5% of the participants had an actionable genotype (n=316 TC, 27.8%, and n=31 CC, 2.7%). The number of participants that discontinued treatment within 3 years (47.1%) was in line with previous findings [10, 49]. We did not find a statistically significant association between *SLCO1B1* and any change in simvastatin or atorvastatin use. The time to stable dose was comparable between included participants regardless of genotype or statin used. In a randomized-controlled trial comparing outcomes between usual care and PGx-informed prescription, Peyser et al. (2018) also did not find a difference in self-reported statin adherence between *SLCO1B1* groups [50]. We did find a statistical significant difference in the starting dose between males and females for simvastatin.

The DCS is a population-based cohort with a relatively long follow-up enabling both cross-sectional and longitudinal comparisons [51], which increases its external validity. Furthermore, we have used routinely collected data of drug dispensing instead of prescriptions through community pharmacies; corrected for known confounders; stratified for type of statin as an effect modifier; and applied multivariate methods to maximize internal validity. However, not all potential covariates could be analyzed, such as thyroid disorder and rheumatoid arthritis, because this data was not available for the DCS [9, 48]. Our major limitation is that we have used proxies to assess the frequency of SRM. Nonetheless, the study has sufficient power through the available sample size. Furthermore, the sample size is comparable to an average Dutch GP practice, so gives a sufficient estimate of the effect for such a primary care practice. Finally, we used a framework for clinical validity and utility, and strived to analyze data beyond ORs which would provide important insights for statistically significant genotype-associations [33].

To assess clinical validity and utility of *SLCO1B1* we suggest a prospective study to monitor actual SRM instead of using proxies, include relevant lifestyle, drug-related, and biological factors.[52, 53] SNPs in other genes alone or in combination with *SLCO1B1* c.521T>C might also influence the risk for SRM, such as the SNP c.421C>A in the *ABCG2* gene and 15389C>T on the CYP3A4 gene [5]. Additional insights might be gathered from an opportunistic genomic screening (OGS) approach, while PGx is not commonly reported in OGS panels [54]. Moreover, the test context in which testing for *SLCO1B1* will be provided in practice, might not be a single-SNP-based approach, but rather a panel-based approach [55]. Therefore, the validity and utility of such an approach will need to be established, including factors such as timing of testing [56]. To study these aspects, outcomes such as the number needed to genotype, and the population attributable fraction of SRM should be studied. Furthermore, the relevant stakeholders should be included in the process, to ensure that outcomes relevant to them are gathered and reported from research [57, 58].

## Conclusion

Our study indicates that *SLCO1B1* c.521T>C screening would not impact relevant factors considering statin use, such as reaching a stable dose sooner and decreasing discontinuation and switches. Participants would not have had a shorter time to establish stable dosing and would not have experienced less SRM. Our findings do suggest that an appropriate method for treatment adherence should be studied, since approximately 47% patients discontinued their treatment, as well as follow-up or review of guidelines for initial statin treatment, because women had a lower starting dose than men. Furthermore, prospective studies focused on ADR need to take into account outcomes for clinical utility, such as predictive characteristics of (a panel of) SNPs and other outcomes relevant to medical doctors, patients, and other stakeholders.

## Data Availability

All data produced in the present study are available upon reasonable request to the authors

## Statements and Declarations

### Availability of data and material (data transparency)

The datasets generated during and/or analysed during the current study are not publicly available as formulated in the informed consent for participants but are available from the corresponding author on reasonable request.

### Code availability

Not applicable

### Author contributions

All authors contributed to the study conception and design. Material preparation, data collection and analysis were performed by TMC Fleur, ME Jansen, T Rigter, PC Souverein, JJ Swen and WMM Verschuren. The first draft of the manuscript was written by ME Jansen and all authors commented on previous versions of the manuscript. All authors read and approved the final manuscript.

### Ethics approval

This research was evaluated by the Medical Ethical Committee of the VU University Medical Center Amsterdam (identifier 2017.074) and the Compliance Committee of the PHARMO Institute.

### Consent to participate

Informed consent was obtained from all individual participants included in the study.

### Consent for publication

Not applicable

## Acknowledgements

We would like to thank R. Baak-Pablo, A. Blokstra, H.M. Hodemaekers, S. Imholz, C. Manoe, E.J.M. Stynenbosch, P.E. Zwart for the laboratory analyses and data preparations in our study. The authors would also like to thank all the healthcare providers contributing information to the PHARMO Database Network.

