## Supplementary Table 1 and 2 for "Predictive value of *SLCO1B1* c.521T>C polymorphism on observed changes in the treatment of 1136 statin-users"

Supplementary table 1. Sample size calculation based on Link *et al.* (2008) in a population of patients receiving 80 mg of simvastatin daily.

| | Controls | Cases | Total |
|---|---|---|---|
| T | 157 | 93 | 250 |
| C | 23 | 77 | 100 |
| Total | 180 | 170 | 350 |
| C allele proportion | 13% | 45% | |
| Needed sample size | 39 | 41 | 80 |

Supplementary table 2. Calculation Hardy Weinberg Equilibrium

| | |
|---|---|
| TT | 789 |
| TC | 316 |
| CC | 31 |

| | |
|---|---|
| TTexp | 789,4445 |
| TCexp | 315,1109 |
| CCexp | 31,44454 |

| | | | |
|---|---|---|---|
| p | 0,833627 | n | 1136 |
| q | 0,166373 | | |

| | | | |
|---|---|---|---|
| Chi-squared | 0,009 | p-value | 0,995488 |

Supplementary table 2. The association between *SLCO1B1* c.521T>C genotype and discontinuation, switch, time to establish a stable dose, and current vs. genotype-based dosing regimen, stratified for statin corrected for different groups of confounders.

| | | **Simvastatin** | **Atorvastatin** |
|---|---|---|---|
| **Dose change** | **Event (%)** | **Adjusted OR (95% CI)** | **Adjusted OR (95% CI)** |
| TT | 30 (4.1) | | |
| TC/CC | 9 (3.0) | 0.46 (0.17-1.23)[a]<br>0.46 (0.17-1.23)[b]<br>0.52 (0.19-1.37)[c] | 1.97 (0.43-9.04)[a]<br>2.52 (0.43-14.95)[b]<br>1.83 (0.42-7.99)[c] |
| Total | 39 (3.4) | | |
| **Change (discontinuation or switch)** | **Event (%)** | **Adjusted OR (95% CI)** | **Adjusted OR (95% CI)** |
| TT | 529 (67.1) | | |
| TC/CC | 223 (64.3) | 0.91 (0.67-1.23)[a]<br>0.90 (0.66-1.21)[b]<br>0.86 (0.64-1.17)[c] | 0.55 (0.28-1.08)[a]<br>0.61 (0.31-1.18)[b]<br>0.62 (0.33-1.17)[c] |
| Total | 752 (66.2) | | |
| **Time to establish stable dosing regimen** | **Median (IQR), days** | **Adjusted OR (95% CI)** | **Adjusted OR (95% CI)** |
| TT | 90 (97) | | |
| TC/CC | 89 (108) | 1.01 (0.86-1.19)[a]<br>1.03 (0.87-1.22)[b]<br>1.05 (0.89-1.24)[c] | 0.87 (0.63-1.20)[a]<br>0.80 (0.57-1.12)[b]<br>0.81 (0.59-1.11)[c] |
| Total | 90 (97) | | |

OR = Odds Ratio; 95% CI = 95% confidence interval

Adjusted for: a. Age, Sex, Diabetes Mellitus, Systolic blood pressure, HDL level; b. BMI, cigarette smoking, physical activity; c. Starting dose, concomitant drug use.

OR = Odds Ratio; 95% CI = 95% confidence interval

Adjusted for: a. Age, Sex, Diabetes Mellitus, Systolic blood pressure, HDL level; b. BMI, cigarette smoking, physical activity; c. Starting dose, concomitant drug use.
